# Shared N417-dependent epitope on the SARS-CoV-2 Omicron, Beta and Delta-plus variants

**DOI:** 10.1101/2022.04.24.22273395

**Authors:** Thandeka Moyo-Gwete, Mashudu Madzivhandila, Nonhlanhla N Mkhize, Prudence Kgagudi, Frances Ayres, Bronwen E Lambson, Nelia P Manamela, Simone I Richardson, Zanele Makhado, Mieke A van der Mescht, Zelda de Beer, Talita Roma de Villiers, Wendy A Burgers, Ntobeko A B Ntusi, Theresa Rossouw, Veronica Ueckermann, Michael T Boswell, Penny L Moore

## Abstract

As SARS-CoV-2 continues to evolve, several variants of concern (VOCs) have arisen which are defined by multiple mutations in their spike proteins. These VOCs have shown variable escape from antibody responses, and have been shown to trigger qualitatively different antibody responses during infection. By studying plasma from individuals infected with either the original D614G, Beta or Delta variants, we show that the Beta and Delta variants elicit antibody responses that are overall more cross-reactive than those triggered by D614G. Patterns of cross-reactivity varied, and the Beta and Delta variants did not elicit cross-reactive responses to each other. However, Beta-elicited plasma was highly cross-reactive against Delta plus (Delta+) which differs from Delta by a single K417N mutation in the receptor binding domain, suggesting the plasma response targets the N417 residue. To probe this further, we isolated monoclonal antibodies from a Beta-infected individual with plasma responses against Beta, Delta+ and Omicron, which all possess the N417 residue. We isolated a N417-dependent antibody, 084-7D, which showed similar neutralization breadth to the plasma. The 084-7D mAb utilized the IGHV3-23*01 germline gene and had similar somatic hypermutations compared to previously described public antibodies which target the 417 residue. Thus, we have identified a novel antibody which targets a shared epitope found on three distinct VOCs, enabling their cross-neutralization. Understanding antibodies targetting escape mutations such as K417N, which repeatedly emerge through convergent evolution in SARS-CoV-2 variants, may aid in the development of next-generation antibody therapeutics and vaccines.

**Importance:** The evolution of SARS-CoV-2 has resulted in variants of concern (VOCs) with distinct spike mutations conferring varying immune escape profiles. These variable mutations also influence the cross-reactivity of the antibody response mounted by individuals infected with each of these variants. This study sought to understand the antibody responses elicited by different SARS-CoV-2 variants, and to define shared epitopes. We show that Beta and Delta infection resulted in antibody responses that were more cross-reactive compared to the original D614G variant, but each with differing patterns of cross-reactivity. We further isolated an antibody from Beta infection, which targeted the N417 site, enabling cross-neutralization of Beta, Delta+ and Omicron, all of which possess this residue. The discovery of antibodies which target escape mutations common to multiple variants highlights conserved epitopes to target in future vaccines and therapeutics.

## Introduction

Since the emergence of SARS-CoV-2 in 2019, several variants of concern (VOCs) and interest (VOIs) have emerged. Many of these variants contain mutations in the spike protein that confer escape from neutralizing antibodies (1). The first neutralization-resistant VOC identified, Beta, contained receptor binding domain (RBD) mutations at positions K417N and E484K, which confer escape from Class I and Class II neutralization antibodies, respectively (2, 3). These mutations have since been observed in multiple variants. One such example is the charge-altering E484K/Q/A mutation, which occurs in several variants including Gamma, the Delta lineage, C.1.2 and A.VOI.V2 (1, 4-6). The E484K mutation is also present in Omicron (first identified in South Africa) which is the most neutralization-resistant variant identified to date, evading both convalescent and vaccine-elicited antibody responses (5, 7-9).

Similarly, the K417N mutation found in the Beta variant has also emerged in other VOC/VOIs throughout the world. This mutation results in a change from a positively charged residue (lysine, K) to an uncharged asparagine (N) residue - a difference large enough to prevent binding of antibodies dependent on this epitope (3). This substitution results in escape from Class I neutralizing antibodies such as the therapeutic monoclonal antibody (mAb), etesevimab (10, 11). The K417N mutation also occurs in a sublineage of Delta, called Delta plus (Delta+), and in the Omicron variant (BA.1) and its sub-lineages (BA.2, BA.3, BA.4 and BA5) (12), indicating that an antibody that targets this site may cross-neutralize these variants.

The high levels of convergent evolution observed globally between variants indicates that at a population level, neutralizing antibodies target similar regions of the SARS-CoV-2 spike protein. This is supported by numerous monoclonal antibody (mAb) isolation studies which identified the RBD as the major target of the neutralizing antibody response, regardless of the infecting variant or type of vaccination (13-17). However, there is substantial evidence that the antibody response towards the RBD differs in fine specificity depending on which variant triggered the infection. The original (D614G) variant triggers antibodies with very low levels of cross-reactivity to the Beta variant (3). Responses elicited by the Beta variant however, exhibit higher levels of cross-reactivity towards the D614G and the Gamma variant (18) but not against the Delta variant (19, 20). Similarly, Delta-elicited responses have low cross-reactivity towards the Beta variant (15, 21). It has therefore been suggested that the Beta and Delta variants took divergent evolutionary pathways towards distinct groupings or “serotypes”, with the D614G original variant being serologically closer to Delta than Beta (19). The Omicron variant elicits predominantly Omicron-specific antibodies, with poor cross-reactivity against Delta and C.1.2 and D614G, and a dramatic 31-fold reduction in activity against Beta (22). Defining the fine specificity of neutralizing antibodies elicited by the different variants is therefore important to determine whether there are common responses targeting divergent variants.

In this study, we compared the cross-reactivity of plasma responses during three separate SARS-CoV-2 waves in South Africa. Each epidemic wave in South Africa was dominated by a distinct variant; first D614G, then Beta, followed by Delta. We first assessed the neutralizing antibody responses elicited by each of the three variants against a panel of circulating VOCs and SARS-CoV-1. We show that Beta and Delta infection elicited more cross-reactive responses than the D614G variant, though the patterns of cross-reactivity were different. Furthermore, Beta-elicited plasma potently neutralized the Delta+ variant compared to the Delta variant, suggesting these cross-reactive antibodies may target the N417 residue, as this is the only spike mutation that differentiates these two variants. To confirm this, we isolated and characterized a mAb from a Beta-infected individual who showed plasma neutralization of Beta, Delta+ and Omicron. Epitope mapping revealed that this mAb, 084-7D, recapitulated much of the plasma breadth and was heavily dependent on a shared N417 epitope. The discovery of cross-reactive mAbs may aid in the development of universal SARS-CoV-2 therapeutic antibodies and inform the design of booster vaccines.

## Results

### Neutralizing responses triggered by variants of concern possess differential patterns of cross-reactivity

We first compared antibody responses in unvaccinated individuals infected by VOCs circulating in three distinct SARS-CoV-2 epidemiological waves in South Africa from May 2020 to July 2021. In each wave, sampling occurred at the time when each respective variant was responsible for at least 90% of infections, and sequencing in a subset of matched nasal swabs confirmed the infecting variant. We investigated how well plasma elicited by D614G, Beta and Delta variants neutralized multiple VOCs, as well as SARS-CoV-1 (**Figure 1A and 1B**). D614G-elicited plasma triggered robust neutralization of the matched D614G spike (autologous neutralization), but showed very low levels of cross-reactivity, with a 12-15-fold decrease in neutralization towards all SARS-CoV-2 VOCs tested and to SARS-CoV-1 (**Figure 1A**). In contrast, as we previously reported, plasma from Beta infections was highly cross-reactive against the D614G variant (2.9 fold reduction) (18) but showed less cross-reactivity against other VOCs, with reductions against Delta, Omicron and SARS-CoV-1 of 11.3-fold, 9.4-fold and 9.3-fold, respectively (**Figure 1A**). We also tested plasma from Delta-infected individuals and found that although autologous titers against the matched spike were very high, there was a 15-41-fold drop in neutralization potency across VOC/VOIs and SARS-CoV-1. The higher level of cross-reactivity for the D614G variant (15-fold drop in potency, compared to Beta (35-fold drop) in Delta-elicited plasma is consistent with what others have reported (19, 21).

**Figure 1:**
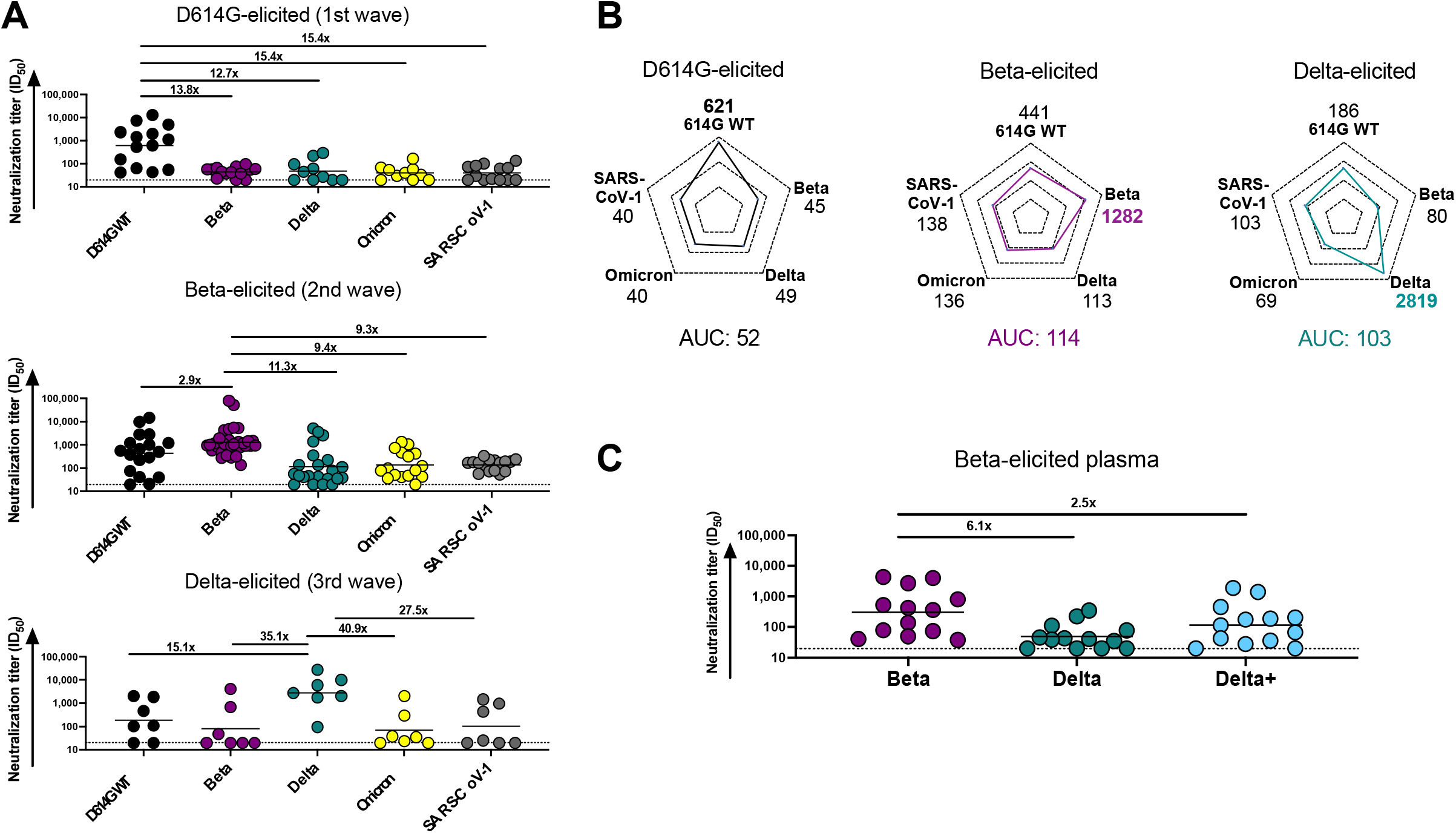
Comparison of plasma cross-reactivity elicited by three distinct SARS-CoV-2 variants. (**A**) Plasma from D614G, Beta and Delta infections during three distinct SARS-CoV-2 waves were tested for neutralization breadth against a range of VOCs using a pseudovirus-based neutralization assay. Fold changes in neutralization are shown above each variant. Plasma neutralization titer is measured as an ID_50_. Black horizontal bars represent geometric means. The threshold of detection for the neutralization assay is ID_50_>20. (**B**) Spider plots were derived from GMTs for plasma triggered by D614G, Beta or Delta against multiple VOCs. The GMTs for each plasma set was normalized against titers to the autologous virus and breadth was expressed as area under the curve. (**C**) Plasma from Beta-infected individuals was tested against the Delta and Delta+ variants. Fold changes in neutralization are shown above each variant. Plasma neutralization titer is measured as an ID_50_. Black horizontal bars represent geometric means. The threshold of detection for the neutralization assay is ID_50_>20.

To get a measure of the degree and patterns of cross-reactivity for each variant, we compared the geometric mean titers (GMTs) of the plasma response elicited by the three variants towards various VOCs and SARS-CoV-1 using a spider plot (**Figure 1B**). Each variant triggered the highest responses against their corresponding autologous virus as expected. Furthermore, the extent of autologous neutralization varied, with Delta triggering particularly high autologous responses (GMT: 2,819), compared to Beta (GMT:1,282) or D614G (GMT:621), likely a consequence of the high viral loads in Delta infections (23). We therefore calculated a single measure of breadth, by assessing the cumulative area under the curve (AUC) value for all variants, normalized relative to the GMT of the autologous virus to account for differential potency of responses (**Figure 1B**). Beta- and Delta-elicited responses were both approximately 2-fold higher (AUC: 114 and 103) than D614G (AUC: 52) (**Figure 1B**), indicating that the neutralizing responses triggered by these two variants were more cross-reactive than those elicited by the early circulating SARS-CoV-2 variant.

Although other sites, such as the RBD Class III antibody binding site, have previously been implicated as targets of the Beta-elicited responses (14) the N417K mutation in the Beta RBD has also been shown to be a dominant target of Beta-elicited plasma (24, 25). To assess whether antibodies to this site accounted for the breadth in Beta-infected individuals, we tested Beta plasma against the Delta+ variant, which differs from Delta only at K417N, the same mutation that is also found in Beta (**Figure 1C**). This mutation is the only shared mutation in the RBD regions of the Beta and Delta+ lineages. Compared to the Beta response towards Delta which showed a significant drop in potency (6.1-fold), there was only a 2.5-fold drop in potency against Delta+ (**Figure 1C**). These data confirm the importance of the N417 residue as a target of Beta-elicited cross-reactive neutralizing antibody responses.

### Isolation and characterization of a SARS-CoV-2 cross-reactive, neutralizing monoclonal antibody from a Beta-infected individual

To assess whether the N417 residue was a target of Beta-elicited mAb responses, we sought to isolate mAbs from a Beta-infected individual who developed potent responses towards the Beta variant and was cross-reactive towards other VOCs. We previously recruited participant SA-01-0084 (084) into our cohort of hospitalized patients infected with the Beta variant (18). This participant was female, below the age of 60, HIV-uninfected, and had their blood drawn two days after a positive SARS-CoV-2 RT-PCR test (**Figure 2A**). Plasma from donor 084 potently neutralized Beta (ID_50_ = 7,613) and Delta+ (ID_50_ = 12,083) pseudoviruses (**Figure 2B**). The 084 plasma also potently neutralized Omicron (ID_50_ = 9,882). However, it did not neutralize the D614G, Delta or SARS-CoV-1 pseudovirus and had weak neutralization against Gamma and A.VOI.V2 (ID_50_<120) (**Figure 2B**). The high cross-reactivity of the plasma towards Delta+ was similar to what we observed with other Beta-elicited plasma (**Figure 1C**).

**Figure 2:**
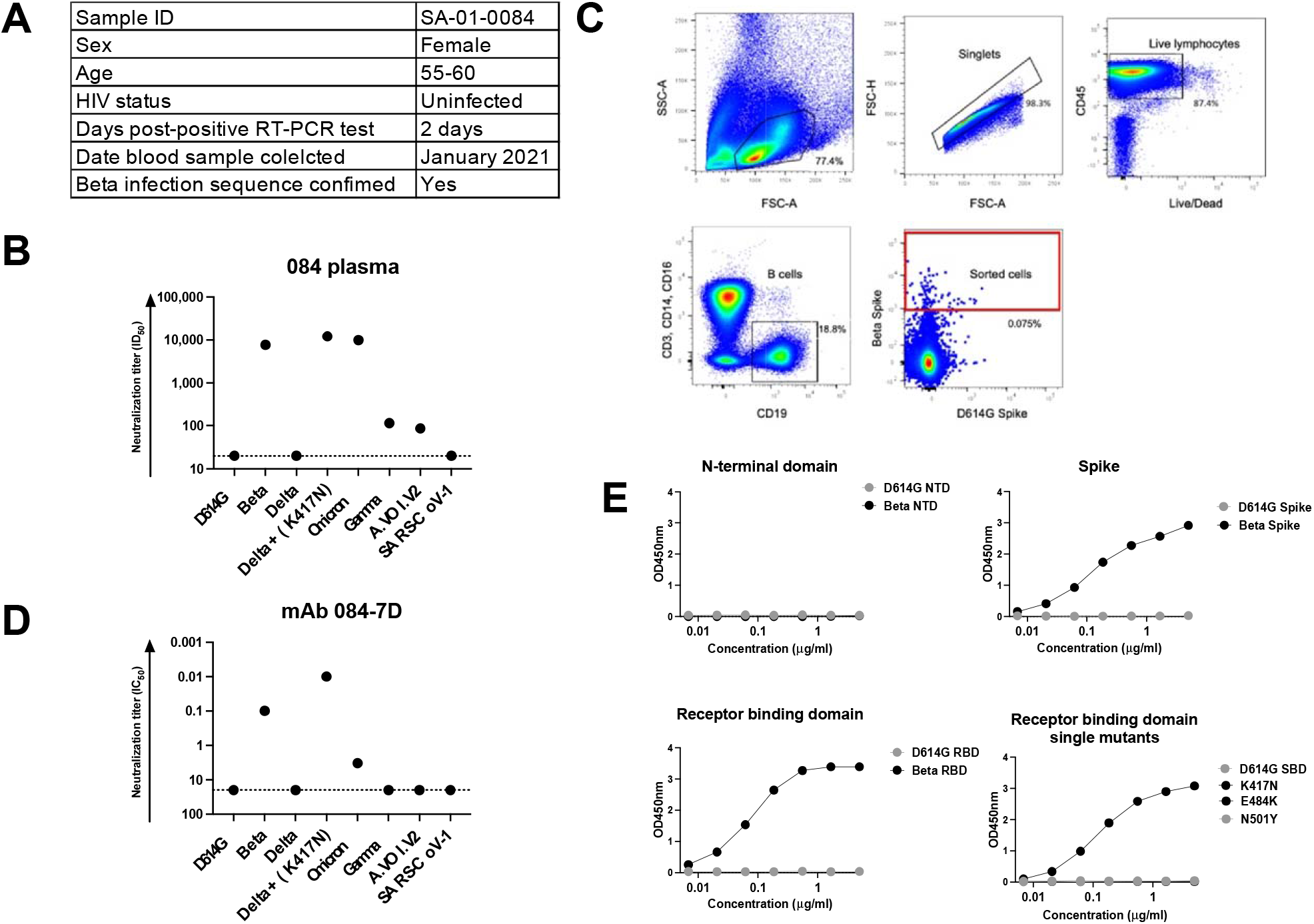
Characterization of plasma and an isolated monoclonal antibody from a Beta-infected individual. (**A**) A sequence-confirmed Beta-infected hospitalized individual, SA-01-0084 (084) was selected for this study. Plasma and peripheral blood mononuclear cells were collected 2 days post-positive PCR for further analysis. (**B**) Plasma from SA-01-0084 was tested for neutralization activity in a pseudovirus-based neutralization assay against a range of VOC/VOIs. Plasma neutralization titer is measured as an ID_50_. The threshold of detection for the neutralization assay is ID_50_>20. (**C**) The gating strategy for the isolation of SARS-CoV-2 spike-specific B cells via single cell sorting is shown. Each dot represents a cell and the dots in the sorted cell red gate represent Beta spike-positive B cells that had CD3-, CD14-, CD16-, CD19+ markers. These cells were single cell sorted into 96 well plates and amplified through antibody gene specific PCR. (**D**) mAb 084-7D was tested for neutralization activity in a pseudovirus-based neutralization assay against the same VOC/VOIs tested with the plasma. mAb neutralization was measured as an IC_50_ (ug/ml). The threshold of detection for the neutralization assay is IC_50_>20 ug/ml (**E**) A SARS-CoV-2 ELISA was used to map the specificity of the 084-7D mAb. The mAb was tested against the D614G and Beta N-terminal domain, spike and RBD antigens, as well as SBD (subdomain 1) proteins mutated to include Beta-specific SBD single mutants, K417N, E484K and N501Y. The starting concentration of the mAb was 5ug/ml. The threshold of detection for the binding assay is OD450nm>0.04.

We performed single cell sorting of the participant’s peripheral blood mononuclear cells (PBMCs). Using Beta and D614G spike proteins as cell sorting baits, we sorted a total of 68 cells, with approximately 58 of the cells being Beta-specific and the rest of the cells being double-positive (**Figure 2C**). We identified an IgG1 mAb, 084-7D, which bound to the Beta spike antigen (**Figure 2**). We tested the functionality of mAb 084-7D by first examining its neutralizing activity against the same VOCs we had tested the plasma against (**Figure 2D**). The 084-7D mAb showed a very similar cross-reactivity pattern to the matched sera, exhibiting potent neutralization of Beta (IC_50_ = 0.10 µg/ml) and Delta+ (IC_50_ = 0.01 µg/ml), though slightly lower activity against Omicron (IC_50_ = 3.31 µg/ml) than was observed in the plasma. The mAb did not neutralize SARS-CoV-1 or the other VOC/VOIs tested (**Figure 2D**). These data indicate that this mAb recapitulated the dominant response in the plasma, though it did not entirely account for Omicron plasma neutralization.

### 084-7D mAb epitope mapping reveals spike target that is highly dependent on the N417 residue

To map the spike target of the mAb, we conducted binding experiments using a SARS-CoV-2-specific enzyme linked immunosorbent assay (ELISA). The antibody bound the full Beta spike but not the D614G spike as expected based on the neutralization assay results (**Figure 2E**). No binding was observed for either the Beta or D614G N-terminal domnain (NTD) proteins, but strong binding was detected towards the Beta RBD (**Fig. 2E**). We expressed single mutants representing the three RBD mutations found in the Beta variant - K417N, E484K and N501Y - in an SBD (subdomain 1) backbone and tested whether the antibody was able to recognize any of the Beta mutations. The 084-7D mAb bound to the K417N mutant but did not bind to the D614G SBD or E484K and N501Y single mutants (**Figure 2E**). Overall, these data suggest that the 084-7D mAb is dependent on the N417 residue present in the Beta RBD.

### mAb 084-7D exhibits Fc effector functionality, particularly against the Beta variant

We have previously shown that, in addition to neutralization, the Beta variant triggers cross-reactive Fc functionality compared to D614G (26). We therefore analyzed the ability of the 084-7D mAb to mediate antibody-dependent cellular phagocytosis (ADCP) and measured the ability of mAb 084-7D to cross-link CD16 and SARS-CoV-2 spike proteins expressed on cells as a proxy for antibody-dependent cellular cytotoxicity (ADCC). Both of these effector functions have been shown to contribute to decreased severity of SARS-CoV-2 infection and are required for optimal protection from infection by monoclonal antibodies (27). We compared mAb 084-7D to control mAbs, P2B-2F6 and CR3022. The 084-7D mAb mediated strong phagocytic activity against the Beta variant (AUC: 1,513) but up to 3-fold weaker activity against the D614G and Omicron variants (AUC: 209 and 468 respectively) (**Figure 3A**). In comparison, P2B-2F6, which targets a Class II RBD epitope, mediated strong ADCP against D614G, but much lower levels against Beta and no activity against Omicron, as expected. The CR3022 mAb showed cross-reactive ADCP to all three variants, as previously reported (22). The 084-7D mAb exhibited ADCC activity against the Beta (AUC: 25,937) and Omicron (AUC: 15,551) variants while no activity was detected for the D614G variant (**Figure 3B**). The P2B-2F6 mAb displayed strong ADCC activity against the D614G variant (AUC:113,631) as expected but no activity towards Beta and Omicron. CR3022 had similar activity across the three variants. Overall, the 084-7D mAb mediated Fc effector functionality with similar patterns of cross-reactivity to that observed in neutralization, exhibiting ADCP and ADCC activity against the Beta variant but reduced activity against Omicron.

**Figure 3:**
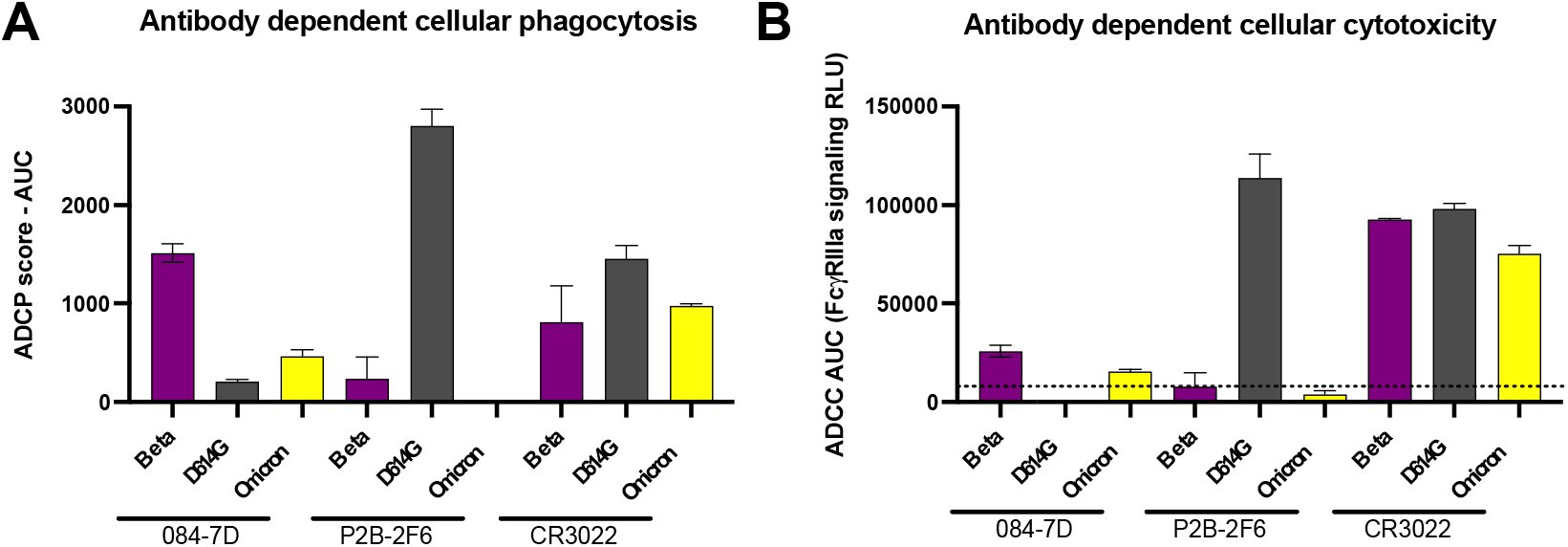
mAb 084-7D displayed antibody dependent cellular phagocytosis (ADCP) and antibody dependent cellular cytotoxicity (ADCC) with cross reactivity towards Beta and Omicron. (**A**) ADCP activity of mAb 084-7D was measured using THP-1 phagocytosis assay and the phagocytosis score shown as an area under the curve (AUC) measure. The threshold of detection for this assay is AUC>0. (**B**) ADCC activity of mAB 094-7D was measured using an infectious ADCC assay. The % killing activity is shown as an AUC measure. The threshold of detection for the ADCC assay is AUC>8000. In both ADCP and ADCC assays P2B-2F6 was used a positive control for D614G activity, CR3022 was used as a positive control for all variants and Palivizumab, an RSV-specific mAb, was used as a negative control. Experiments were conducted in duplicate and error bars represent the mean with standard deviation of two experiments.

### 084-7D is related to a broad and potent N417-dependent antibody isolated from early pandemic infection

We next investigated the genetics of mAb 084-7D to determine whether it could be classified amongst other RBD-targeting mAbs based on germline gene usage and epitope targeting (**Figure 4**). The 084-7D antibody was isolated as an IgG1, IgA1 and IgM but was cloned and tested as an IgG1 for this study (**Figure 1A**). The mAb uses the 3-23*01 variable heavy (VH) chain gene, exhibiting 5.9% somatic hypermutation in the variable gene region, despite being isolated early in infection (**Figure 4A**). The 1-5*03 variable kappa (VK) chain gene is used by this mAb, and we observed less somatic hypermutation in the VK gene (2.9%) compared to the heavy chain. We compared the 084-7D mAb to a previously reported SARS-CoV-2 RBD-specific antibody, CAB-A17, that utilizes the VH3-53*01 germline gene, which is similar to VH3-23*01 with 91% identity. The CAB-A17 lineage developed VOC cross-neutralization through interactions with the N417 residue, suggesting similar epitope targeting compared to mAb 084-7D (28).

**Figure 4:**
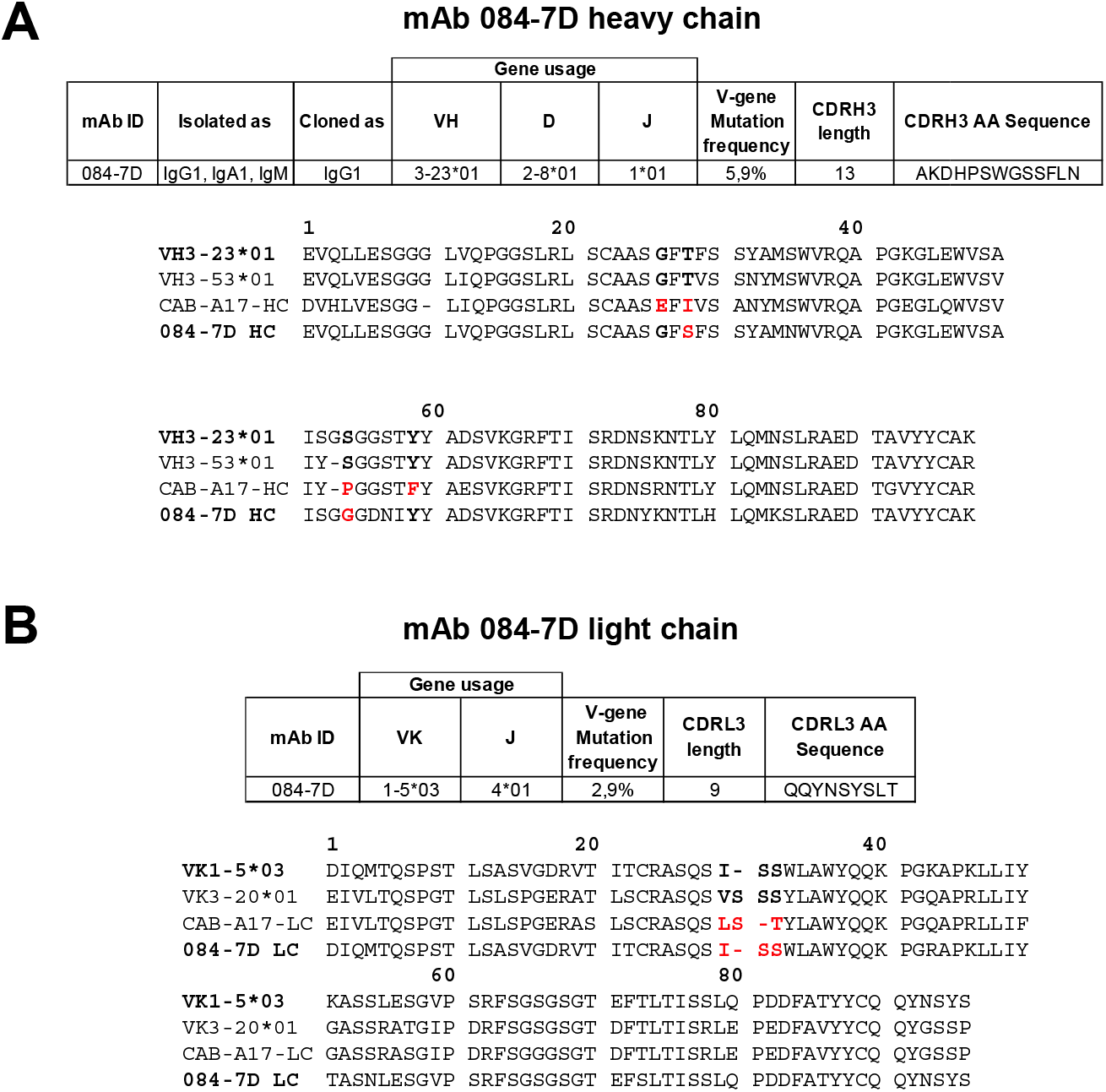
Genetic analysis of mAb 084-7D compared to a similar N417-dependent mAb. (**A**) Heavy chain and (**B**) light chain analysis of mAb 087-7D was conducted using IMGT/V-QUEST (https://www.imgt.org/IMGT_vquest/analysis). A multiple sequence alignment of the germline gene and mAb sequences for 084-7D and CAB-A17 was generated using ClustalW (https://www.genome.jp/tools-bin/clustalw). Key positions implicated in the development of breadth in the CAB-A17 mAb are shown in bold and mutations from germline gene residues in these regions are shown in red.

Sheward *et al*. identified four key mutations in the heavy chain of the broad mAb, CAB-A17, which likely contributed to the increased breadth observed: G26E, T28I, S53P and Y58F (Kabat numbering). Alignment of the VH3-23*01 (used by 084-7D), VH3-53*01 (used by CAB-A17), CAB-A17 and 084-7D heavy chain sequences (truncated at the CDRH3 region) showed that the 084-7D mAb had mutations at two of the same key residues, T28S and S53G, although the amino acid changes differed from the CAB-A17 mAb mutations (**Figure 4A**).

Furthermore, the light chain of Class I K417-targeting antibodies has been shown to play a role in epitope interactions through the CDRL1 ^28^VSSS^31^-motif (Kabat numbering) (13, 28). We noted that the 084-7D mAb possessed a germline encoded ISS-motif, which is similar to the CAB-A17 mAb which also had a deletion in this region, resulting in an LST-motif (**Figure 4B**). Altogether, these data show that the 084-7D mAb is similar to a highly broad and potent mAb which targets a similar epitope.

## Discussion

In this study, we investigated the cross-reactive potential of plasma elicited by three different variants that circulated in South Africa during three sequential waves of infection - the D614G, Beta and Delta variants. Antibodies generated by the Beta and Delta variants exhibited higher levels of cross-reactivity as compared to those triggered by the original D614G variant confirming that SARS-CoV-2 variants elicit varying antibody responses, potentially targeting different epitopes on the spike protein. Secondly, we used a mAb isolated from a Beta-infected donor to define a N417-dependent neutralizing antibody epitope shared between the Beta, Delta+ and Omicron variants. This mAb exhibited similar genetic features to a potent and broad N417-dependent antibody, suggesting that this site may be commonly targeted by SARS-CoV-2 cross-reactive mAbs.

The K417N mutation has been implicated in immune escape from Delta-elicited plasma and this may explain the lack of cross-reactivity between Beta and Delta antibody responses (15). In line with this, we found that although Delta+ and Delta spike sequences only differ by one amino acid (N417 in Delta+ and K417 in Delta), antibodies elicited by the Beta variant showed cross-reactivity against the Delta+ VOI but very little cross-reactivity towards the Delta VOC. Our findings suggest that the Beta-elicited plasma responses we tested were preferentially targeting an epitope that was dependent on the N417 residue that is found in the Beta, Delta+ and Omicron VOCs. Our data confirms findings by other groups who have shown that the N417-directed response accounts for a large proportion of the Beta-elicited response in some individuals (24, 25).

Here, we isolated an N417-dependent mAb from a Beta-infected individual which neutralized and performed Fc effector function against the Beta, Delta+ and Omicron VOCs. The 084-7D mAb utilizes the VH3-23*01 germline gene, and members of this class of mAbs have previously been shown to target the RBD (29, 30). VH3 gene families are well-documented in targeting this site, including Class I antibodies which bind to the K417-residue (13, 29). The VH3 group of antibodies have been classed as public antibodies suggesting that they can be easily elicited in SARS-CoV-2 infection and vaccination (29, 31, 32). Similar to the 084-7D mAb, low levels of somatic hypermutation are needed for most VH3 SARS-CoV-2 antibodies to neutralize their target, suggesting that germline-encoded sequences, such as in the CDRH2 and CDRL2, play a sufficient role in epitope recognition (13). These characteristics of mAb 084-7D make it attractive to pursue as a vaccine target.

Although many aspects of neutralization are germline encoded, increased somatic hypermutation enhances the maturation of breadth to SARS-CoV-2 VOCs (33), as for other pathogens (34, 35). As mAb 084-7D was isolated from blood drawn only two days after testing positive for SARS-CoV-2, this antibody was isolated too early to have accumulated substantial somatic hypermutation. Plasma from donor 084 collected at the same timepoint was highly potent against Omicron, but the 084-7D mAb displayed less potency against this variant. This suggests that there may be other antibodies in the plasma which are cross-reactive against Omicron.

The lack of neutralization and Fc effector of mAb 084-7D against Omicron compared to the Beta and Delta+ variants may be impacted by the fact that the Beta and Delta+ variants each possess three mutations in their RBD regions, including the K417N mutations, whilst Omicron possesses at least 17 mutations in that region (5). The numerous mutations clustered around the Y501 residue in Omicron may have an impact on mAb 084-7D interactions as this region has been shown to be important for Class I antibody CDRL1 interactions (29). Although our data focused on the interaction between this mAb and the N417 residue, the complete epitope-paratope interaction must be investigated through the determination of a high-resolution structure. This will delineate the role of other residues in creating the full epitope of this mAb to decipher its relatively lower levels of potency towards Omicron.

A recently described mAb, CAB-A17, shares similar genetic and epitope-dependent traits to mAb 084-7D (28). However, mAb CAB-A17 has substantially more breadth across VOCs and is also more potent against Omicron than the 084-7D mAb (28). Exploring the differences between the two mAbs may suggest a mechanism required to achieve high levels of breadth and potency in this class of antibody. The CAB-A17 mAb contains key mutations that resulted in increased breadth towards the Omicron variant - G26E, T28I, S53P and Y58F. The 084-7D mAb contains 2/4 of these mutations although the residue changes differ - T28S and S53G. The changes in mAb CAB-A17 at these sites are much more substantial, especially the serine to proline change at position 53 that may alter the conformation of that region and therefore affect epitope binding. These differences may contribute to the disparity in breadth and potency between these two mAbs.

In addition to their distinct epitope-paratope interactions, the angle of approach is likely different between the two mAbs. CAB-A17 makes interactions with the N417 residue through the Y33 residue on the heavy chain (28). mAb 084-7D possesses an alanine at that position (A33) that would likely be unable to make strong contacts with the asparagine at 417 due to its shorter side chain. Therefore, it is plausible that mAb 084-7D interacts with the N417 residue using an alternative contact site(s) or binds through a different angle of approach. It is important to note that the evolution of the CAB-A17 mAb took place over seven months while 084-7D was isolated from acute infection. Tracking the evolution of mAb 084-7D will reveal whether somatic hypermutation led to increased potency and breadth and whether this maturation mimicked the pathway used by CAB-A17 or followed an independent route.

The identification of another novel N417-dependent mAb that can neutralize various VOC/VOIs indicates that this may be a good target for a cross-reactive response. The CAB-A17 mAb was isolated earlier in the pandemic, presumably as a result of D614G infection, while the 084-7D mAb was isolated from Beta infection, again highlighting the universal targeting of this epitope by different variants. Despite the divergence of variants into “serotypes” or “clusters”, we show that the antibody response towards different variants can converge on shared epitopes. Finding mAbs that target conserved sites across variants will likely aid in the development of therapeutic mAb cocktails that can treat SARS-CoV-2 infection regardless of the infecting variant.

## Materials and Methods

### Cohort description

Plasma samples from the first SARS-CoV-2 wave (D614G-infected) were obtained from a previously described cohort across various sites in South Africa prior to September 2020 (3). Second wave samples (Beta-infected) were obtained from a cohort of patients admitted to Groote Schuur Hospital, Cape Town in December 2020 - January 2021 (18). Third wave samples (Delta-infected) were obtained from the Steve Biko Academic Hospital, Tshwane from patients admitted in July 2021. In all waves, samples were collected when more than 90% of SARS-CoV-2 cases in South Africa were caused by the respective variants. Sequence confirmation was only available for a subset of samples but all the samples that were sequenced corresponded to the appropriate variant for that wave. All samples were from HIV-uninfected individuals who were above 18 years of age and provided consent. Ethical clearance was obtained for each cohort from Human Research Ethics Committees from the University of Pretoria (247/2020) and University of Cape Town (R021/2020).

### SARS-CoV-2 antigen design and expression

The pseudovirus SARS-CoV-2 Wuhan-1 spike was mutated using the QuikChange Lightning Site-Directed Mutagenesis kit (Agilent Technologies) and NEBuilder HiFi DNA Assembly Master Mix (NEB) into the 614G WT (D614G wild-type), Beta (L18F, D80A, D215G, 242-244del, K417N, E484K, N501Y, D614G and A701V), Delta (T19R, 156-157del, R158G, L452R, T478K, D614G, P681R and D950N), Delta plus (Delta+) (T19R, 156-157del, R158G, K417N, L452R, T478K, D614G, P681R and D950N), Gamma (L18F, T20N, P26S, D138Y, R190S, K417T, E484K, N501Y, D614G, H655Y, T1027I, V1176F) and Omicron (A67V, Δ69-70, T95I, G142D, Δ143-145, Δ211, L212I, 214EPE, G339D, S371L, S373P, S375F, K417N, N440K, G446S, S477N, T478K, E484A, Q493R, G496S, Q498R, N501Y, Y505H, T547K, D614G, H655Y, N679K, P681H, N764K, D796Y, N856K, Q954H, N969K, L981F). The SARS-CoV-1 spike was obtained from Genscript. Pseudoviruses were prepared as previously described (3). Briefly, human embryonic kidney (HEK) 293T cells were co-transfected with the SARS-CoV-2 spike plasmid of interest together with a firefly luciferase encoding lentivirus backbone plasmid for 72 hours. Culture supernatants were filter-sterilized and stored at −70°C.

For soluble proteins, the SARS-CoV-2 D614G spike was obtained from Jason McLellan (University of Texas) and the SARS-CoV-2 RBD WT from Florian Krammer. The full spike proteins were mutated to produce the 614G WT (D614G) and Beta (L18F, D80A, D215G, 242-244del, K417N, E484K, N501Y, D614G and A701V), Delta (T19R, 156-157del, R158G, L452R, T478K, D614G, P681R and D950N, A.VOI.V2 (D80Y, DEL144, I210N, DELN211, D215G, R246M, DEL LAL247-249,W258L, R346K, T478R, E484K, H655Y, P681H, Q957H) and Omicron (BA.1) (A67V, Δ69-70, T95I, G142D, Δ143-145, Δ211, L212I, 214EPE, G339D, S371L, S373P, S375F, K417N, N440K, G446S, S477N, T478K, E484A, Q493R, G496S, Q498R, N501Y, Y505H, T547K, D614G, H655Y, N679K, P681H, N764K, D796Y, N856K, Q954H, N969K, L981F) spike proteins. SARS-CoV-2 subdomain 1 (SBD) protein (residues: 321-591) and N-terminal domain (NTD) protein (residues: 14-304) were designed in-house, ordered from Genscript and cloned into a mammalian cell expression vector. Mutagenesis was used to produce SBD Beta (K417N, E484K or N501Y) SBD K417N, SBD E484K, SBD N501Y single mutants and NTD Beta (L18F, D80A, D215G, 242-244 del). Proteins were expressed in human embryonic kidney HEK293F cells for 6 days, at 37°C, 70% humidity and 10% CO_2_. Purification was carried out using a nickel-charged resin followed by size-exclusion chromatography. Pure protein fractions were collected and frozen at -80°C until further use.

### SARS-CoV-2 monoclonal antibody isolation

Cryopreserved PBMCs from a SARS-CoV-2-infected participant, SA-01-0084, were stained for SARS-CoV-2-specific B-cell markers before single cell sorting. B-cells marked as CD3-, CD14-, CD16-, CD19+ and Aqua Vital Dye-negative were selected. Biotinylated SARS-CoV-2 Beta and D614G spike antigens were labelled with PE and AF647, respectively, through the biotin-streptavidin interaction. Viable cells that bound the Beta and/or D614G spikes were single-cell sorted into 96 well plates and heavy and light chain genes were PCR amplified.

### Single cell PCR amplification of heavy and light chain variable genes

Genes encoding IGHV and IGLV chains were amplified from sorted cells by reverse real-time and nested PCR. Reverse transcription was performed using Superscript III reverse transcriptase (Invitrogen) and random hexamer primers (36, 37). Following cDNA synthesis VH, Vκ, and Vλ antibody genes were amplified as previously described (38, 39). Determination of gene usage was done using IMGT/V-QUEST (https://www.imgt.org/IMGT_vquest/analysis).

### Pseudovirus-based neutralization assay

Heat-inactivated plasma samples from COVID-19 convalescent donors or mAbs were incubated with the SARS-CoV-2 pseudoviruses for 1 hour at 37°C, 5% CO2. Subsequently, 1×10^4^ HEK 293T cells engineered to over-express ACE-2, provided by Michael Farzan, were added and incubated at 37°C, 5% C02 for 72 hours upon which the luminescence of the luciferase gene was measured. Monoclonal antibodies CB6, CA1 and CR3022 were used as controls.

### SARS-CoV-2 enzyme linked immunosorbent assay (ELISA)

96-well high-binding plates with 2 µg/ml of the respective antigen were incubated overnight at 4°C. After blocking with 5% milk powder in 1x PBS and 0.05% Tween-20, mAbs at 5 µg/ml starting concentration was serially diluted and incubated at room temperature for 1.5 hours. Subsequent washing was followed by the addition of an anti-human horseradish peroxidase secondary antibody diluted to 1:3000 and the plates were incubated for a further 1 hour. TMB substrate (ThermoFisher Scientific) was added, followed by 1M H_2_SO_4_ to stop the reaction, with absorbance of the reaction measured at an optical density of 450nm. mAbs CR3022, P2B-2FB and Palivizumab served as controls.

### Antibody-dependent cellular cytotoxicity (ADCC) assay

The ability of the mAb to cross-link between FcγRIIIa (CD16) and spike expressed on cells was used as a proxy for ADCC. To express cell-surface spike, HEK 293T cells were transfected with SARS-CoV-2 614G WT, Beta or Omicron expressing plasmids, with incubation at 37°C over 2 days. Spike expressing cells were incubated with 100 µg/ml of 084-7D or control mAbs titrated four times 1 in 5 in RPMI media, 10% FBS, 1% penicillin-streptomycin for an hour at 37°C. Following this, Jurkat-Lucia™ NFAT-CD16 cells (Invitrogen) were added to the reaction, and incubated for a further 24 hours at 37°C with 10% CO_2_. Signal was read on a luminometer by adding 20µl of supernatant and 50µl QUANTI-Luc secreted luciferase to white 96-well plates. CR3022, P2B-2F6 and Palivizumab served as controls and data across spikes were normalized using CR3022.

### Antibody-dependent cellular phagocytosis (ADCP) assay

Biotinylated SARS-CoV-2 614G WT, Beta and Omicron spike proteins were coated onto beads by incubating fluorescent neutravidin beads with 10 µg/ml antibody for 2 hours as described elsewhere (22). Overnight incubation was carried out with monocytic THP-1 cells, followed by analysis on the FACSAria II (BD Biosciences). The percentage of engulfed fluorescent beads within the THP-1 cells multiplied by the geometric mean was calculated and a final phagocytosis score was determined by subtracting the fluorescence score of the no antibody control. CR3022, P2B-2F6 and Palivizumab served as controls.

## Data Availability

All data produced in the present study are available upon reasonable request to the authors

## Acknowledgements

We acknowledge the participants who volunteered for this study. The parental soluble SARS-CoV-2 spike was provided by J McLellan (University of Texas), parental pseudovirus plasmids by Drs E Landais and D Sok (IAVI) and HEK 293T-ACE-2 cells were provided by Dr Michael Farzan (Scripps Research).

